# Sociodemographic factors associated with Public knowledge of dementia in a Cuban population

**DOI:** 10.1101/2021.06.04.21258359

**Authors:** Zoylen Fernández-Fleites, Yunier Broche-Pérez, Claire Eccleston, Elizabeth Jiménez-Puig, Evelyn Fernández Castillo

**Author notes:** **Corresponding author information:** Prof. Zoylen Fernández-Fleites, MSc., Universidad Central “Marta Abreu” de Las Villas, Psychology Department Carretera de Camajuaní, Km 5½ Santa Clara, Villa Clara, Cuba, 54830. **Ethical approval statement:** The data was obtained following the regulations of the ethics committee of the Department of Psychology of the Universidad Central “Marta Abreu” de Las Villas and in accordance with the Declaration of Helsinki. **Financial disclosure:** The authors involved in this research communication do not have any relationships with other people or organizations that could inappropriately influence (bias) the findings.

## Abstract

**Objectives:** To explore knowledge and beliefs among a cross-section of the Cuban adult population with regard to dementia risk factors and to determine the demographic variables related with it.

**Study design:** A cross-sectional survey was carried out on 1004 Cubans.

**Methods:** The survey measured the importance placed on dementia, risk reduction knowledge and the actions to prevent it. Logistic regression was undertaken to identify variables associated with knowledge.

**Results:** Most respondents (47.5%) believe that dementia risk reduction should start before age of 40. Cognitive stimulation and physical activities were selected with major frequency. Being older than 48 years, having previous contact with dementia and university education increases the probability of having healthy lifestyles.

**Conclusions:** The exploration of demographic variables allows the prediction of likelihood to know about or have positive beliefs in relation to dementia. They should be contemplated into strategies for dementia prevention in Cuban population.

## Introduction

International organizations estimate that a new dementia diagnosis is given every three seconds.[1] The number of cases will be more marked in low and middle income countries, which will experience more than two third of all cases. Latin America constitutes one of the regions that will be more affected in next 20 years, with an increase of dementia diagnosis of 146 percent.[2]

In this context, dementia prevention is of greater relevance. The evidence supports that around 40% worldwide dementia can be attributed to 12 risk factors. Those 12 risk factors are potentially modifiable during life-course to dementia prevention, and requires both public health programmes and individually tailored interventions. [3]For example, a healthy lifestyle and controlling cardiovascular risk factors may be able to reduce a third of all cases of dementia diagnosed.[4-6] In Latin American (LA) countries the dementia prevention potential is greater than in high-income countries. According to a recent study, the overall weighted population attributable fractions (PAF) for potentially modifiable risk factors for dementia in Latin American countries is around 55 percent.[7]

This is particularly important given that Cuba is the Latin American country with the highest percentage of older adults. In Cuba life expectancy is over 78 years and 30% of the Cuban population is expected to be aged over 60 by 2030.[8] In general terms, the increase in life expectancy has been associated also with a sustained increase in non-communicable diseases, including neurodegenerative disorders, like dementia.[9]

The prevalence of dementia in Cuba in people over 65 has been estimated at 10.8%, ranking third in the Spanish-speaking Caribbean region behind the Dominican Republic (11.7%) and Puerto Rico (11.6%).[10] These results have been obtained under the umbrella of 10/66 studies, a multinational initiative aimed to provide a detailed scientific evidence for improving the health and quality of life of older people in low and middle income countries, including Cuba.[11]

Also as part of the 10/66 initiative, a recent study carried out in the Cuban population identified for the first time the main risk factors of incident dementia in Cuban older adults and explores how these factors change with age.[12]

The study was conducted with a sample of 1,846 Cuban participants who are part of the 10/66 study. The participants were stratified into two age groups (65-74 years and 75+ years). In the youngest group (65-74 years), the risk factors that showed a positive association with the incidence of dementia in the participants were the presence of stroke, ischemic heart problems, depression, and physical activity. In the oldest group (75+ years) the variables positively associated with the development of dementia were smoking status, physical activity, and fish consumption.[12]

We agree with the study authors that these results open a path towards the development of prevention strategies that allow reducing the incidence of dementia in the Cuban population. In this scenario, it is important to explore dementia knowledge in the general population. [13] However, most of the research in this field has been conducted in high income nations.[14-19], despite the higher prevalence of dementia in regions with middle and low income.[20]

Recently, a systematic review was conducted with the aim of synthesizing the general public’s perceptions of dementia in Latin America region.[21] The research identified only six studies about this topic in the region; all of them from Brazil. According the authors, there was evidence of a limited to moderate knowledge of dementia, though a significant minority also had negative or stigmatizing attitudes. Only higher levels of education were consistently associated with better attitudes and knowledge of dementia in the region.[21]

In the case of Cuba, there is only one study aimed at exploring the knowledge about dementias in the general population.[22] The authors of the study surveyed a total of 391 people aged between 18 and 96 years. As the main result it was reported that a total of 64.5% of participants believed that the risk of dementia could be reduced, and 60% that the appropriate time to begin prevention measures is after the age of 40.

However, the study has several limitations that prevent generalizing the results to the Cuban population. Among the main limitations of the research are the size of the sample and the purely descriptive nature of its conclusions. For example, it would be interesting to know how the level of knowledge is associated with sociodemographic and clinical variables, which would allow the development of specific actions aimed at modifying risk factors.

Studies that explore the level of knowledge about dementias in the general population are especially useful within the framework of national strategies aimed at the prevention of dementias and the care of patients diagnosed with this syndrome.

In the case of Cuba, Dementia’s National Strategy has developed as part of the answer to the international call toward the need for global strategies for better dementia diagnosis, treatments interventions and prevention actions. The Cuban strategy defend the need for dementia prevention and healthy aging promotion.[23] An important step for the development of prevention actions is to explore the level of knowledge that exists in the general Cuban population about the risk factors related to the development of dementia.

In this scenario, the main objective of the present study is to explore knowledge and beliefs among a cross-section of Cuban adult population with regard to the risk factors that may lead to dementia, and the actions that may be taken to prevent it. Additionally, it aims to explore the relationships between demographic variables and dementia knowledge among the Cuban population.

## Methods

### Material and subjects

A cross-sectional survey was carried out between January 2019 and February 2020, with a total of 1004 participants. All Cuban (18 years old or more, and to consent to participate) were able to participate. Sample was random in the central region of Cuba, in urban areas. The objectives of investigation were explained to each potential participant, and the survey was carried on with those that offered informed consent. The dementia knowledge includes dementia information based in evidence, with relevance for general public and health staff, [24, 25] and a pathway to better treatment and care of the person with the condition and also to prevention. [26] According to that, items were taken from previous survey used by Smith, Ali [14] The survey has a total of six questions. Question 1 registers personal information (gender, age, educational level and previous contact with a person with dementia). Question 2 explores personal health concerns. Question 3 asks whether the respondent believes that the risk of dementia can be reduced (this question has five response options, ranging from ‘‘it cannot be reduced’’ to ‘‘it can be reduced’’). Question 4 explores the knowledge of actions that can reduce the risk of experiencing dementia (this question is open-ended in order to evaluate participants’ actual knowledge without introducing response bias). The open-ended responses were categorized using the groups used by Smith, Ali [14] as a basis. The participants’ beliefs about the activities that could be taken to reduce the risk of dementia were explored in question 5 (open-ended too), in order to explore those activities incorporated into participants’ lifestyle. Finally, Question 6 enquires about the age at which preventive measures should be taken, in the respondent’s opinion. The survey were conducted by trained interviewers.

### Statistical methods

In bivariate analysis, the Chi-squared statistics were used to compare dementia beliefs and knowledge between different strata of respondents. The independent variables in these analysis were gender (male/female); age group (18-27 years, 28-47 years, more than 48 years, as a continuity of Broche-Pérez, Fernández-Fleites [22] research); educational level (primary level, secondary level, high school, university); and previous contact with a person with dementia (yes, no). Backwards step-wise logistic regression modelling was undertaken [27] with those variables where bivariate relationships were observed, in order to identify those associated with dementia beliefs and knowledge. A maximum *p*-value of 0.10 was adopted for the retention of variables at each step of the model, in order to promote direct comparison with the work of Smith, Ali [14]. Analysis was made using SPSS v21.0. For this sample size, a power analysis was running (post hoc) using the G*Power software (version 3.1.9.2) [28]. Considering the sample size, the analysis (two-tailed) showed a power of 0.91.

### Procedures and Ethics

The research protocol was approve by ethics committee from Psychology Department of Universidad Central “Marta Abreu” de Las Villas. The investigation was developed according to the World Medical Association Declaration of Helsinki.

## Results

### Sample characteristics

In total, 1, 004 interviews were carried out. The characteristics of respondents are shown in table 1. The mean age was 41.4 years (SD 17.7) and most respondents were aged over 48 years old (38.4%). A higher proportion were women (55.4%). Just over 30% were educated at university level and the majority were employed (65.9%). More than 50% had had prior contact with people with dementia.

**Table 1.** Characteristics of survey respondents (N=1004)

| **Characteristics** | **Value(%)** |
| --- | --- |
| Mean age (SD) | 41.40(17.7) |
| Age groups |  |
| < 27 years | 311(31.0) |
| 28-47 years | 307(30.6) |
| >48 years | 386(38.4) |
| Gender* |  |
| Women | 556(55.4) |
| Men | 446(44.4) |
| Education Level** |  |
| Primary school | 20(2.0) |
| Secondary School | 129(12.8) |
| Pre-university studies | 502(50.0) |
| University | 325(32.4) |
| Work status*** |  |
| Employed | 662(65.9) |
| Retired | 172(17.1) |
| Student | 70(7.0) |
| Home duties | 30(3.0) |
| Contact with people with dementia**** |  |
| Yes | 591(58.9) |
| No | 396(39.4) |

*Notes.* \*(2 people did not respond), \*\* (28 people did not respond), \*\*\* (70 people did not respond), \*\*\*\* (17 people did not respond)

#### Prevalence of knowledge and beliefs about dementia risk reduction

As shown in table 2, only 32% of survey respondents rated dementia as one of the three most important health issues; as a result dementia was placed in a fourth level after brain tumor (40.3%), heart disease (37.7%), and breast cancer (37.2%). Dementia was considered one of the main health issues in 38% of people older than 48 years, and in 27% of participants in remaining groups.

**Table 2.** Beliefs about dementia as a health issue, dementia risk reduction and the best age to adopt dementia risk reduction behaviours

|  | Health issue | Dementia risk reduction |  |  |  | Best age to start prevention |  |  |  |
| --- | --- | --- | --- | --- | --- | --- | --- | --- | --- |
|  |  | Definitely it can | I think it can | I don't know | I think it can't | Definitely it can't | <40 years | 40-59 years | >60 years |
| Total* | 31.7 | 62.4 | 2.4 | 25.5 | 1.1 | 8.7 | 47.5 | 35.0 | 17.3 |
| Gender |  |  |  |  |  |  |  |  |  |
| Women | 31.8 | 63.1 | 2.9 | 25.5 | 1.3 | 7.2 | 47.9 | 36.9 | 15.1 |
| Men | 32.2 | 61.2 | 1.8 | 25.6 | 0.9 | 10.5 | 47.4 | 32.6 | 20.0 |
| Age group |  |  |  |  |  |  |  |  |  |
| < 27 years | 27.3** | 59.5* | 3.2* | 27.0* | 2.3* | 8.0* | 59.4*** | 26.1*** | 14.5*** |
| 27-48 years | 27.7** | 61.9* | 2.3* | 29.0* | 0.0* | 6.8* | 51.3*** | 35.9*** | 12.7*** |
| > 48 years | 38.3** | 65.0* | 1.8* | 21.5* | 1.0* | 10.6* | 35.2*** | 41.5*** | 23.3*** |
| Education level |  |  |  |  |  |  |  |  |  |
| Primary school | 45.0* | 40.0 | 0.0 | 25.0 | 5.0 | 30.0 | 20.0*** | 40.0*** | 40.0*** |
| Secondary School | 29.5* | 48.1 | 3.1 | 34.9 | 0.8 | 13.2 | 34.1*** | 38.8*** | 27.1*** |
| Pre-university studies | 27.7* | 62.9 | 3.4 | 24.1 | 1.4 | 8.2 | 49.0*** | 34.2*** | 16.8*** |
| University | 36.9* | 70.8 | 0.6 | 22.2 | 0.6 | 5.8 | 51.4*** | 35.4*** | 13.2*** |
| Contact with people with dementia |  |  |  |  |  |  |  |  |  |
| Yes | 37.2*** | 66.8** | 1.5** | 22.8** | 1.0** | 7.8** | 52.4*** | 33.2*** | 14.4*** |
| No | 23.7*** | 56.6** | 3.0** | 29.0** | 1.3** | 10.1** | 40.0*** | 38.0*** | 22.0*** |

Notes. Chi-square\*p < 0.05; \*\*p < 0.01; \*\*\*p < 0.001; all values are in percent; Health issue = dementia rated as one of the three most important health issues)

Also 62% of participants strongly agreed that dementia could be reduced, with a quarter of respondents indicating that they didn’t know (Table 2). Two thirds (67%) of those with prior contact with dementia agreed that dementia risk reduction was possible. The age group considered best for the beginning of dementia prevention varied across the sample (Table 2). Most respondents (47.5%) believe that dementia risk reduction should start before age of 40. Participants older than 48 years considered ages between 40 and 59 years as the best time to start adopting risk reducing behaviours, with nearly a quarter believing the best time was over 60 years of age.

Knowledge of behaviors that are beneficial for dementia risk reduction and dementia risk reduction behaviors incorporated into participants’ lifestyles are shown in table 3. The action that respondents most often identified as beneficial for lowering dementia risk was mental activity (63%), followed by physical activity (47%), healthy diet (43%), low alcohol consumption (37%) and social activity (33%) (Table 3). Behaviors like taking vitamins, reducing cholesterol and leisure cognitive activities were not often identified.

**Table 3.**
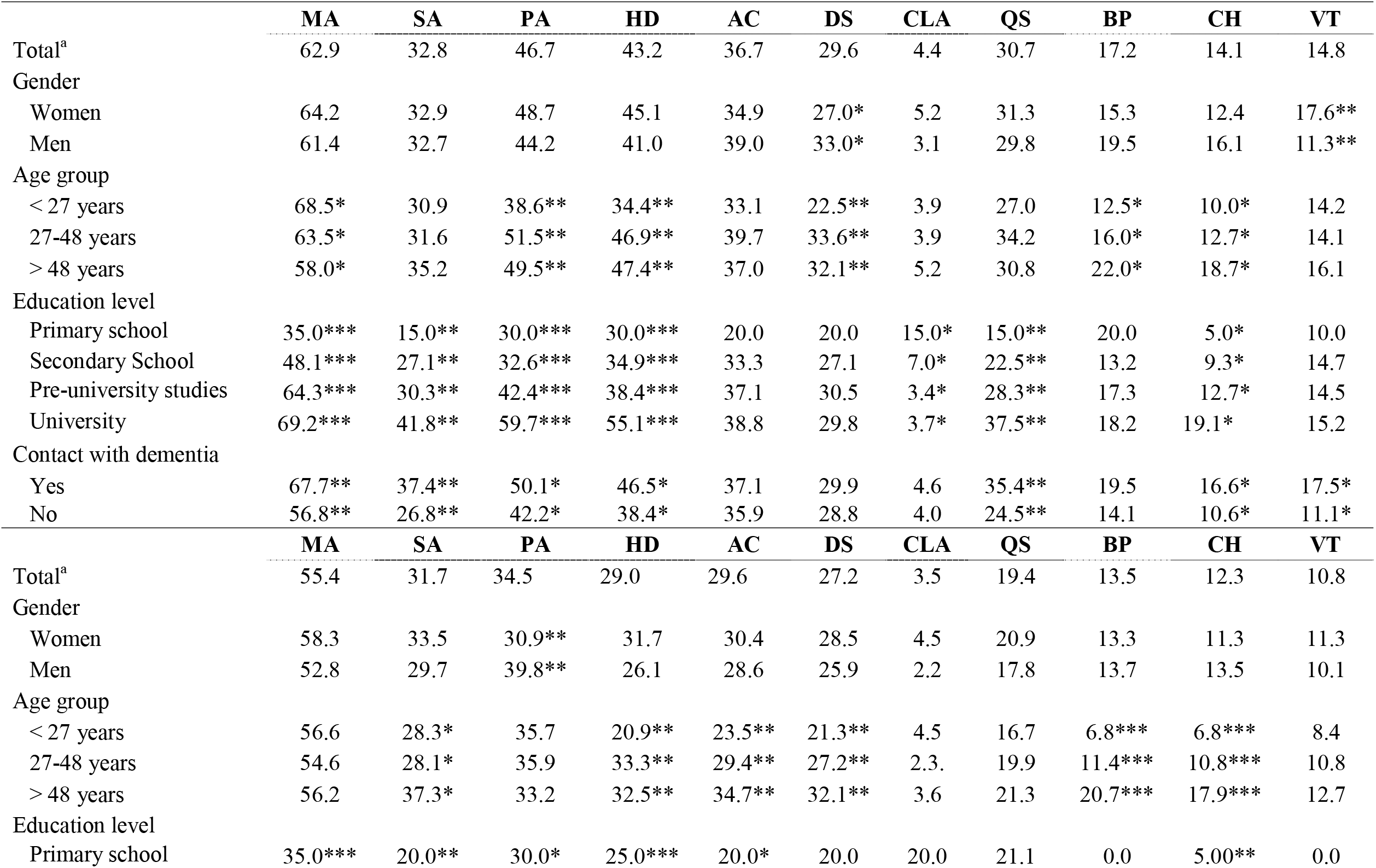
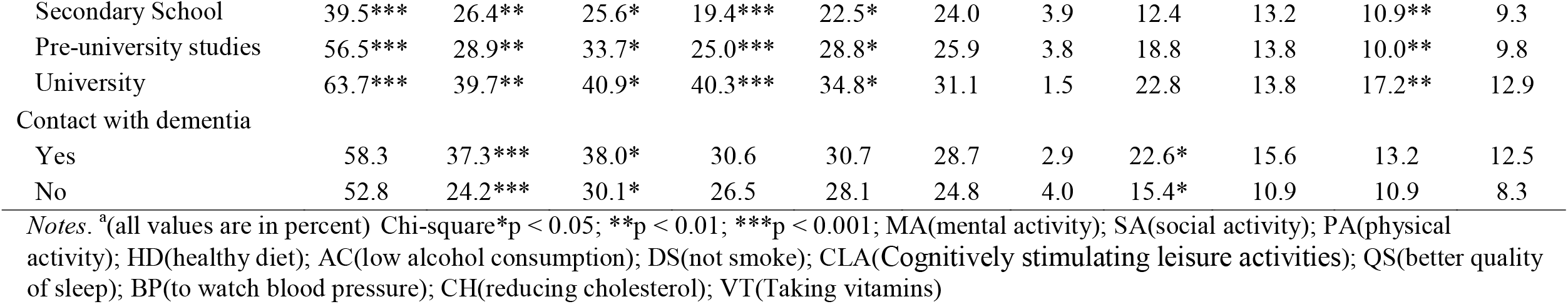
(a) Knowledge of behaviors that are beneficial for dementia risk reduction and b) dementia risk reduction behaviors incorporated into participants’ lifestyles

|  | MA | SA | PA | HD | AC | DS | CLA | QS | BP | CH | VT |
| --- | --- | --- | --- | --- | --- | --- | --- | --- | --- | --- | --- |
| Total<sup>a</sup> | 62.9 | 32.8 | 46.7 | 43.2 | 36.7 | 29.6 | 4.4 | 30.7 | 17.2 | 14.1 | 14.8 |
| Gender |  |  |  |  |  |  |  |  |  |  |  |
| Women | 64.2 | 32.9 | 48.7 | 45.1 | 34.9 | 27.0* | 5.2 | 31.3 | 15.3 | 12.4 | 17.6** |
| Men | 61.4 | 32.7 | 44.2 | 41.0 | 39.0 | 33.0* | 3.1 | 29.8 | 19.5 | 16.1 | 11.3** |
| Age group |  |  |  |  |  |  |  |  |  |  |  |
| < 27 years | 68.5* | 30.9 | 38.6** | 34.4** | 33.1 | 22.5** | 3.9 | 27.0 | 12.5* | 10.0* | 14.2 |
| 27-48 years | 63.5* | 31.6 | 51.5** | 46.9** | 39.7 | 33.6** | 3.9 | 34.2 | 16.0* | 12.7* | 14.1 |
| > 48 years | 58.0* | 35.2 | 49.5** | 47.4** | 37.0 | 32.1** | 5.2 | 30.8 | 22.0* | 18.7* | 16.1 |
| Education level |  |  |  |  |  |  |  |  |  |  |  |
| Primary school | 35.0*** | 15.0** | 30.0*** | 30.0*** | 20.0 | 20.0 | 15.0* | 15.0** | 20.0 | 5.0* | 10.0 |
| Secondary School | 48.1*** | 27.1** | 32.6*** | 34.9*** | 33.3 | 27.1 | 7.0* | 22.5** | 13.2 | 9.3* | 14.7 |
| Pre-university studies | 64.3*** | 30.3** | 42.4*** | 38.4*** | 37.1 | 30.5 | 3.4* | 28.3** | 17.3 | 12.7* | 14.5 |
| University | 69.2*** | 41.8** | 59.7*** | 55.1*** | 38.8 | 29.8 | 3.7* | 37.5** | 18.2 | 19.1* | 15.2 |
| Contact with dementia |  |  |  |  |  |  |  |  |  |  |  |
| Yes | 67.7** | 37.4** | 50.1* | 46.5* | 37.1 | 29.9 | 4.6 | 35.4** | 19.5 | 16.6* | 17.5* |
| No | 56.8** | 26.8** | 42.2* | 38.4* | 35.9 | 28.8 | 4.0 | 24.5** | 14.1 | 10.6* | 11.1* |
|  | MA | SA | PA | HD | AC | DS | CLA | QS | BP | CH | VT |
| Total<sup>a</sup> | 55.4 | 31.7 | 34.5 | 29.0 | 29.6 | 27.2 | 3.5 | 19.4 | 13.5 | 12.3 | 10.8 |
| Gender |  |  |  |  |  |  |  |  |  |  |  |
| Women | 58.3 | 33.5 | 30.9** | 31.7 | 30.4 | 28.5 | 4.5 | 20.9 | 13.3 | 11.3 | 11.3 |
| Men | 52.8 | 29.7 | 39.8** | 26.1 | 28.6 | 25.9 | 2.2 | 17.8 | 13.7 | 13.5 | 10.1 |
| Age group |  |  |  |  |  |  |  |  |  |  |  |
| < 27 years | 56.6 | 28.3* | 35.7 | 20.9** | 23.5** | 21.3** | 4.5 | 16.7 | 6.8*** | 6.8*** | 8.4 |
| 27-48 years | 54.6 | 28.1* | 35.9 | 33.3** | 29.4** | 27.2** | 2.3. | 19.9 | 11.4*** | 10.8*** | 10.8 |
| > 48 years | 56.2 | 37.3* | 33.2 | 32.5** | 34.7** | 32.1** | 3.6 | 21.3 | 20.7*** | 17.9*** | 12.7 |
| Education level |  |  |  |  |  |  |  |  |  |  |  |
| Primary school | 35.0*** | 20.0** | 30.0* | 25.0*** | 20.0* | 20.0 | 20.0 | 21.1 | 0.0 | 5.00** | 0.0 |
| Secondary School | 39.5*** | 26.4** | 25.6* | 19.4*** | 22.5* | 24.0 | 3.9 | 12.4 | 13.2 | 10.9** | 9.3 |
| Pre-university studies | 56.5*** | 28.9** | 33.7* | 25.0*** | 28.8* | 25.9 | 3.8 | 18.8 | 13.8 | 10.0** | 9.8 |
| University | 63.7*** | 39.7** | 40.9* | 40.3*** | 34.8* | 31.1 | 1.5 | 22.8 | 13.8 | 17.2** | 12.9 |
| Contact with dementia |  |  |  |  |  |  |  |  |  |  |  |
| Yes | 58.3 | 37.3*** | 38.0* | 30.6 | 30.7 | 28.7 | 2.9 | 22.6* | 15.6 | 13.2 | 12.5 |
| No | 52.8 | 24.2*** | 30.1* | 26.5 | 28.1 | 24.8 | 4.0 | 15.4* | 10.9 | 10.9 | 8.3 |

*Notes.* <sup>a</sup>(all values are in percent) Chi-square\*p < 0.05; \*\*p < 0.01; \*\*\*p < 0.001; MA(mental activity); SA(social activity); PA(physical activity); HD(healthy diet); AC(low alcohol consumption); DS(not smoke); CLA(Cognitively stimulating leisure activities); QS(better quality of sleep); BP(to watch blood pressure); CH(reducing cholesterol); VT(Taking vitamins)

Mental activity was the activity most commonly considered important by participants of both genders (men 64%, women 61%). Also women mentioned the importance of some lifestyle factors, such as a healthy diet more frequently than men, and the opposite occurs for other factors such as low alcohol consumption and non-smoking. People older than 27 years old are more likely to correctly select risk reducing activities than younger people are; even for those activities less often selected by the general sample such as reducing cholesterol and cognitive leisure activities.

The risk reduction behaviors more commonly reported as part of the daily life of participants were mental activity, physical activity, social activity and low alcohol consumption (Table 3). However, other activities such as watching blood pressure and cholesterol and cognitively stimulating leisure activities, were often not included in participant’s lifestyles. A higher proportion of women reported participating in social activity (34%) and having a healthy diet (32%) in comparison to men (30% and 26% respectively), while higher proportions of men stated that they participate in physical activity (40%).

### Multivariable analysis of factors associated with dementia beliefs and knowledge

Table 4 shows demographic and social factors associated with dementia risk reduction beliefs and knowledge and dementia risk reduction behaviors incorporated in participants’ lifestyle. Respondents aged 48 and over were significantly more likely to consider dementia as a very important health issue (1.53) than those in the younger group (Table 4). In relation to this, people with previous dementia contact are 1.54 times more likely than those without contact to recognize that dementia is preventable. Also, people aged 48 and over were significantly more likely to report physical activity (1.67) and a healthy diet (1.70) than younger people, which means that being 48 years old or more increases by 1.7 the likelihood of identifying the impact of those activities in dementia prevention. Participants educated at university level were more likely to report mental (3.52), social (3.70) and physical (3.72) activity compared to those with primary schooling only. Not having prior contact with people with dementia also decreases the likelihood to select mental activity (in 0.66) and social activity (in 0.63) in comparison to those with previous contact.

**Table 4.**
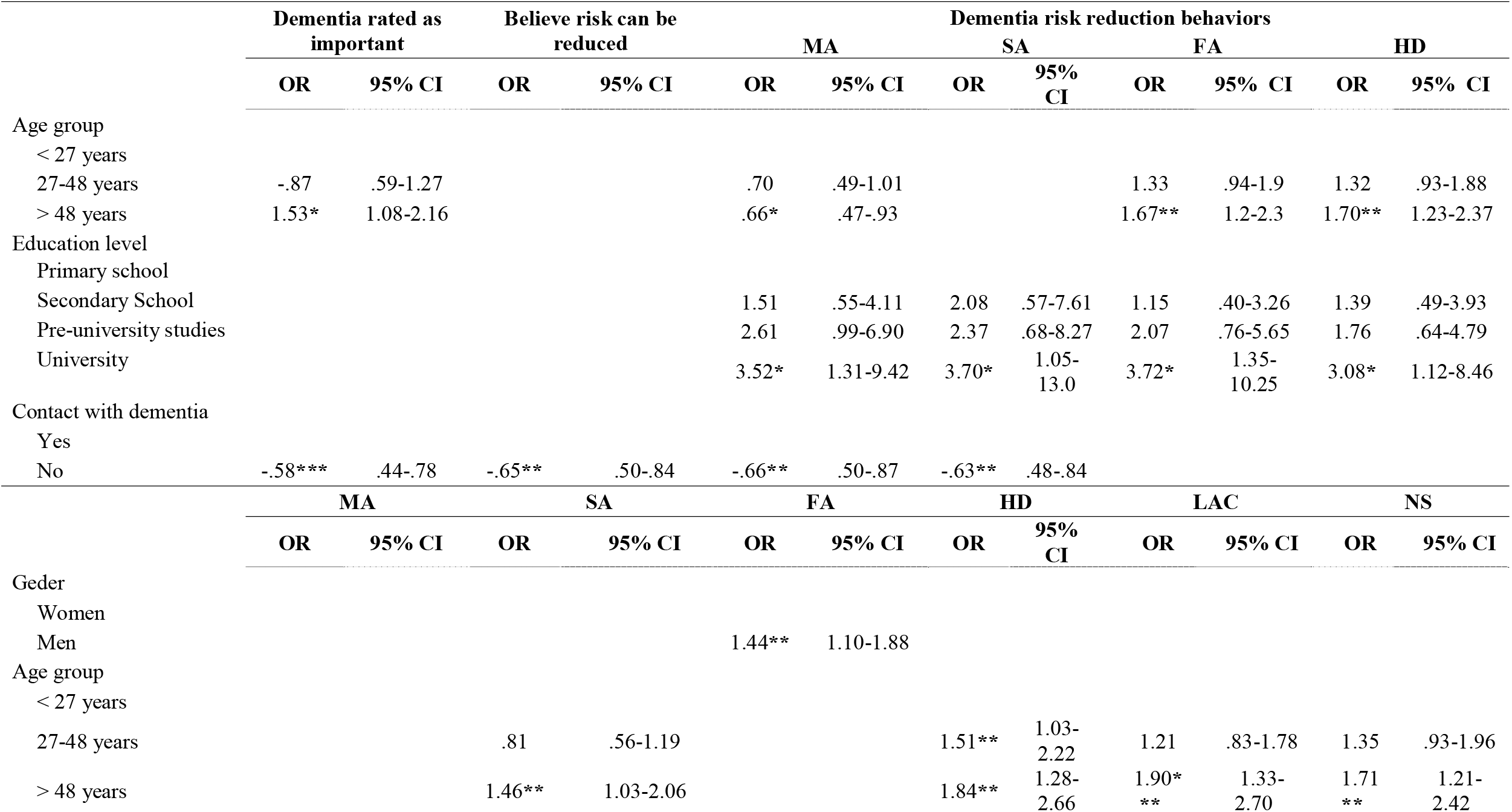
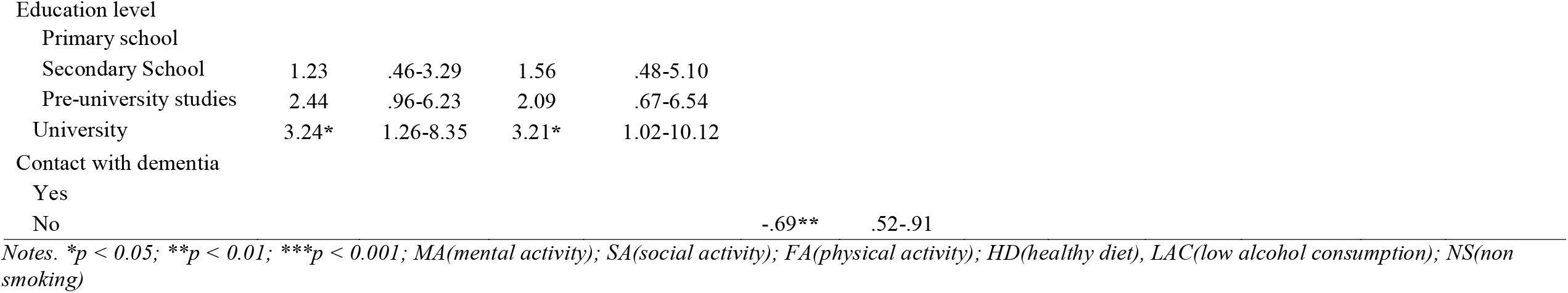
Demographic and social factors associated with (1) dementia risk reduction beliefs and knowledge and (2) dementia risk reduction behaviors incorporated in participants’ lifestyle

As table 4 shows, being 48 years old or more (1.46) and having a university level of education (3.21) increase the likelihood of participants integrating social activity into their lifestyle to prevent dementia.

Table 5 shows the odds of identifying a specific age at which it is best to start dementia prevention, according to those related variables. People without prior dementia contact are 1.59 times higher than people with prior experience of dementia (-.63) to consider 40-59 years as the best time to start prevention, rather than at a younger age. This indicates that people who have had prior contact with people with dementia know about early dementia prevention and that they are optimistic about its possibilities. According to the age groups sampled, those younger than 27 years (-.36) and those between 28-47 years old (-.64) have lower odds of selecting 40-59 as the best time to start prevention compared with the selection of a younger age. It means that both older and younger people believe the appropriate time to begin dementia prevention is this middle age period.

**Table 5.** Demographic and social factors associated with best age group to start adopting risk reducing behaviours

|  | 95% CI |  |  |
| --- | --- | --- | --- |
|  | Inf. | OR | Sup. |
| 40-59 years old vs. younger than 40 years <sup>a</sup> |  |  |  |
| Age group |  |  |  |
| < 27 years | .25 | -.36*** | .54 |
| 27-48 years | .44 | -.64* | .94 |
| > 48 years |  |  |  |
| Contact with dementia |  |  |  |
| Yes | .46 | -.63** | .87 |
| No |  |  |  |
| after 60 years old vs. before 40 years old <sup>a</sup> |  |  |  |
| Age group |  |  |  |
| < 27 years | .22 | -.36** | .58 |
| 27-48 years | .28 | -.46** | .75 |
| > 48 years |  |  |  |
| Education level |  |  |  |
| Primary school | 1.18 | 4.55* | 17.59 |
| Secondary School | 1.35 | 2.51** | 4.65 |
| Pre-university studies | .87 | 1.40 | 2.23 |
| University |  |  |  |
| Contact with dementia |  |  |  |
| Yes | .33 | -.49*** | .73 |
| No |  |  |  |
| Notes. <sup>a</sup>(reference category); *p < 0.05; **p < 0.01; ***p < 0.001 |  |  |  |

## Discussion

The main objective of the present study was to explore knowledge and beliefs among a cross-section of Cuban adults with regard to the risk factors that may lead to dementia, and the actions that may be taken to prevent it. Additionally, we have examined demographic variables related to dementia knowledge in the Cuban population.

The study shows that many Cubans do not recognize dementia as a health priority, although the results for this question are better to those from some other countries.[14, 29] The result is, however, similar to that reported by Russo, Bartolini [30], who found that Argentinians list dementia as the third most worrying condition (preceded by cancer and cerebrovascular disease), with 15% considering it of most concern.

Understanding the extent of knowledge about the possibility of reducing dementia risk is crucial to develop actions to increase the general public’s risk perception. Our results offered values that are in the scope of the results obtained in other studies. In a study conducted in Australia, 72% of participants stated that they were certain that the risk of experiencing dementia could be lowered [31], while 17% of North Americans indicated the opposite, that dementia risk cannot be reduced.[29] Cations, Radisic [32] describe how more than a half of the surveyed population described dementia as a non-preventable condition, therefore our results (only 8.7% of the sample) evidence optimism regarding this matter. These result is probably related to Cuban implementation of the Global Plan of Action for dementia.[23]

In relation to this, people with previous dementia contact are 1.54 times more likely than those without contact to recognize that dementia is preventable. Even though there are some studies with health professionals (doctors and nurses) which indicate there is an inadequate knowledge about the risk factors of the disease, and even of the cognitive symptoms associated with pathology [33-36], there is a link between experience of people living with dementia and knowledge. For example, Eccleston, Doherty [37] show dementia knowledge can be positively related to previous learning about dementia from various types of exposure to the condition (having family members and/or working with people living with the condition).

### Knowledge about the age of start for dementia prevention

More than a half of the younger group of participants considered ages younger than 40 years as suitable to start addressing dementia prevention. Nearly a quarter of older participants (48 years and over) considered ages over 60 years as appropriate to start prevention, a belief which could lead to a failure to adopt preventive behaviours at the appropriate time. Recent evidence points to ages between 40-59 years old as suitable to begin dementia prevention and address important risk factors.[38-40] Offering appropriate information about this could therefore have a notable impact on the risk reduction of developing the disease.

Participants older than 48 years old, without university level education and without prior contact with dementia, show more likelihood of selecting later age groups as the best for starting to address dementia prevention, in comparison to reference groups, and they constitute potential risk groups that should receive particular educational efforts.

### Knowledge about dementia risk reduction behaviors

Unlike other contexts where less than 50% of the general public consider that there are actions to reduce dementia risk [17], in our sample more than 60% thought it possible. Participants considered cognitive stimulation to be the most beneficial activity to reduce risk, followed by a healthy lifestyle (physical activity, healthy diet, low alcohol consumption). With regard to this matter, Livingston, Huntley [3] recent review found no evidence of generalized cognition improvement from specific cognitive interventions, although the domain trained might improve. Also, authors explain that few high quality studies and no long-term high quality evidence about prevention of dementia currently exists. Additionally, there is enough evidence to include alcohol consumption and physical inactivity as 2 of 12 modifiable risk factors for dementia.[3] It is clear, therefore, that participants have an inadequate understanding of the best behavioural change to make in order to reduce the risk of dementia.

Also of particular interest is that women mentioned the importance of some lifestyle factors, such as a healthy diet more frequently than men, and the opposite occurs for other factors such as low alcohol consumption and non-smoking. According to Smith, Ali [14], these differences can be a reflection of the social construction of gender and its impact on health beliefs. Differences by gender have been also noted by other authors in the area of mental health. For example, Lee, Hwang [41] identified a statistically significant difference in the level of mental health attitude between genders, with males having a significantly more negative attitude than females.

Some behaviors such as taking vitamins and better quality of sleep showed low frequency of recall in our participants, which constitutes a positive result given recent studies describing the lack of evidence about the role of this element in dementia prevention [3].

Many healthy behaviors showed a significant association with demographic variables, except gender. This variable showed significance only predicting knowledge about benefits from non-smoking for dementia prevention. This result coincides with reports in Ireland by Glynn, Shelley [17], but differs from data described by Smith, Ali [14], where gender predicts the probability of mentioning the importance of mental activity, social activity, and healthy diet. On the other hand, being more than 48 years old, with university education and prior contact with dementia increase the probability of knowing the future benefit of healthy behaviors. Similar results have been reported in previous studies.[14, 17, 19] It is important to highlight that even though the knowledge of dementia risk factors is not guaranteed for behavioral change, knowledge constitutes a basic component for dementia prevention.[22]

### Healthy behaviors in participants’ lifestyle

In our study, participants of 48 years old and more, with university education, and prior contact with dementia were more likely to implement healthy lifestyles in comparison to participants without these characteristics. Additionally, in our study we find a dissociation between “knowing” and “doing” related to the implementation of healthy behaviors in dementia prevention. A low frequency of preventive actions was confirmed just as in previous studies in the Cuban population (e.g. Broche-Pérez, Fernández-Fleites [22], and this was despite knowledge held by the population about beneficial behaviors. Additionally, implementing a single healthy behavior was considered by most participants to be sufficient to prevent dementia. Therefore, future actions should be redirected to promote the idea of more effective dementia prevention via reduction of several risk factors, as pointed out by several researchers in this field.[42-44]

## Conclusions

The growing agreement about the role of lifestyle changes in reducing the risk of dementia has focused recent research into public knowledge about these topics. It important to highlight that knowledge about dementia might have other benefits too. For example, knowing that it is not a normal part of aging may encourage people to seek a diagnosis. This survey found, however, that knowledge and beliefs in Cuban population are not currently high enough for effective reduction of population risk of dementia. The study demonstrates the need to give greater attention to public education about dementia prevention in order to promote the benefits of a healthy lifestyle and management of cardio-vascular risks. The results presented constitute a baseline for measurement of changes in public knowledge and beliefs, and allow the identification of population risk groups to guide educational actions, particularly in persons without prior contact with dementia.

### Limitations and future directions

This research is not without limitations. First, authors recognize issues with using a backwards step-wise model. Some other authors describe that stepwise methods have no value for theory testing. However, in our case, backward stepwise method were used in a situation in which no previous research exists, only one exploratory precedent [22] as recommended by Field [45]. Also, authors considered important to explore knowledge about some others recent described risks factors such as diabetes, traumatic brain injury, and air pollution, which consequently could theoretically be prevented or delayed dementia [3]. It also would be important to explore some other psychosocial and psychological predictors like the role of caregiver or principal caregiver, according to the importance of previous contact with dementia for disease knowledge. [37] In addition, another element to consider in the study is the lack of control of the participants by province or region of the country. In this sense, future exploration of sociocultural components and the health policy in each region or province of the country can offer significant information for the results and new programs to promote cognitive health in dementias.

## Data Availability

The data associated with the study can be requested directly from the corresponding author of the manuscript.

